# Serum lipidome associates with neuroimaging features in patients with traumatic brain injury

**DOI:** 10.1101/2023.03.14.23287262

**Authors:** Ilias Thomas, Virginia F. J. Newcombe, Alex M. Dickens, Sophie Richter, Jussi P. Posti, Andrew I. R. Maas, Olli Tenovuo, Tuulia Hyötyläinen, András Büki, David K. Menon, Matej Orešič, CENTER-TBI MR subgroup Participants and Investigators

## Abstract

Acute traumatic brain injury (TBI) is associated with substantial metabolic abnormalities, both centrally and in the periphery. We have previously reported extensive changes in the circulating metabolome resulting from TBI, including changes proportional to disease severity and associated with patient outcomes. The observed metabolome changes in TBI likely reflect several pathophysiological mechanisms supporting the concept that TBI is a systemic disease after the primary injury. However, one of the main metabolic changes we have observed following a TBI are changes in lipids, including the structural lipids that are known to be present in the myelin in the brain.

Here, we conducted a study to investigate the relationship between traumatic microstructural changes in white matter seen on magnetic resonance imaging (MRI) and quantitative lipidomic changes in the blood in a subset of patients with TBI recruited to the MRI sub-study of the Collaborative European NeuroTrauma Effectiveness Research in TBI (CENTER-TBI) study.

In total, there were 103 patients who had both a magnetic resonance imaging (MRI) scan and serum samples available for analysis. From serum, 201 known lipids were quantified. Diffusion tensor fitting generated fractional anisotropy (FA) and mean diffusivity (MD) maps for the MRI scans, in addition to volumetric data. Association matrices and partial correlation networks were built to elucidate the connections between the lipid groups and the maps.

We found that there are distinct directions of associations between the neuroimage data (FA and MD sets) and the concentrations of circulating lipids after injury. The FA and MD values were in inverse relationship with the severity of TBI (higher MD values, lower FA). We also observed that the lipid associations to FA and MD show different metabolic signatures. Lysophosphatidylcholines (LPC) associate mostly with FA while sphingomyelins (SM) associate with MD. Only phosphatidylcholines(PC) have strong associations with both as well as with the volumetric data. Finally, we found that the lipid changes are not associated with the number of regions with abnormalities.

In conclusion, we have identified groups of lipids which assocate with specific MRI imaging metrics following TBI. There appears to be consistent patterns of lipid changes associating with the specific microstructure changes in the CNS white matter. There is also a pattern of lipids with regional specficity, suggesting that blood-based lipidomics may provide an insight into the underlying disease mechanisms in TBI.

## Introduction

Traumatic brain injury (TBI) affects over 50 million people worldwide every year^1,2^. Despite being so common, the dynamic pathophysiology and determinants of outcome trajectories remain poorly understood; TBI has been described as the most complex disease in the most complex organ^3^. This is particularly true when attempting to understand the changes that may occur in circulating metabolites after TBI. Understanding these metabolic abnormalities is critical, since such knowledge might allow us to design and evaluate novel therapies. Indeed, we already know that TBI is associated with substantial metabolic abnormalities, which have been mainly demonstrated in humans using positron emission tomography^4,5^ and magnetic resonance spectroscopy^6,7^. However, these techniques are expensive and logistically demanding, and are thus not suitable for large studies or broad clinical implementation.

Metabolomic and lipidomic analysis of systemic blood provides one convenient approach to address this issue. We have previously reported extensive changes in the circulating metabolome resulting from TBI, including changes proportional to disease severity and associated with patient outcomes^8-10^. The observed metabolome changes in TBI likely reflect several pathophysiological mechanisms supporting the concept that TBI is a systemic disease. However, one of the main metabolic changes we have observed following a TBI are changes in lipids^10^, that are known to be present in the myelin in the brain. Myelin is rich in lipids which constitute approximately 80% of the dry weight of myelin, and this makes changes in lipid profile a prime target for characterizing damage to the brain. Previously, the correlations that we have demonstrated between metabolomics and lipidomic changes and computed tomography (CT) are limited, since CT does not characterize microstructural abnormalities. Furthermore, CT does not provide any detailed information on the state of the myelin in the central nervous system (CNS) and therefore, associating the lipid changes with damage to the myelin is challenging especially since this is an important driver of outcome^11^.

Magnetic resonance imaging (MRI) has the potential to characterize microstructural damage and improve our understanding of the pathophysiology underlying different metabolomic profiles after TBI. This technique provides researchers with a window into the axonal and myelin health in the CNS. In particular, advanced quantitative MRI, including diffusion tensor imaging (DTI), have been shown to be sensitive to injury after TBI, and has been associated with the severity of injury and outcome^12-14^. DTI in particular is able to detect microstructural damage especially in the white matter., which may provide insights into the pathophysiology of injuries^15^. Combining this information with lipid measures from the blood will allow us to better understand how changes in the myelin in the CNS can impact the host lipidome profile. Previous studies have demonstrated that circulating lipids can be utilized as a biomarker for microvascular disease^16^. In assessing the acute phase of TBI and the aftermath thereafter, lipidomic profiling may provide more information about the integrity of brain connectivity and, more generally, brain health.

Here we report a study to investigate the relationship between traumatic microstructural changes in the brain seen on MRI and quantitative lipidomic changes in the blood in a subset of patients recruited to the MRI sub-study of the Collaborative European NeuroTrauma Effectiveness Research in TBI (CENTER-TBI) study.

## Materials and methods

### Study participants

The CENTER-TBI study recruited 4509 patients from 18 European countries and Israel (https://www.center-tbi.eu/, registered at clinicaltrials.gov NCT02210221)^17^. The CENTER-TBI database contains data from 65 centers whose data were collected between December 19, 2014, and December 17, 2017. Ethical approval was obtained by each site in accordance with their local regulations (for details see https://www.center-tbi.eu/project/ethical-approval). Informed consent was obtained from all study participants or their legal representatives/relatives according to the local regulations of each center. Clinical data was accessed via the Neurobot platform (RRID/SCR_017004, core data, version 3.0; International Neuroinformatics Coordinating Facility; released November 24, 2020).

Patients were included in the analysis for this study if they were aged ≥18 years, had blood samples taken within 24 hours of injury and had an MRI scan performed within four weeks of injury. For patients who had multiple MRI scans, the earliest one was used. Imaging data from healthy controls were used to harmonize imaging data and detect MRI abnormalities. In total data from 104 healthy controls were used (Table 1), and more are provided in the following sections. All severities of TBI were included. Patients who had significant preexisting neurological disorders were excluded.

**Table 1.** Patient and control demographic characteristics.

|  | TBI patients | Healthy controls |
| --- | --- | --- |
| Number of subjects | 103 | 104 |
| Age (interquartile range) | 43 (28.5-58) | 39.5 (28.5-58) |
| Sex | 77M/26F | 61M/42F (1 missing) |
| Baseline GCS (interquartile range) | 15 (10.75-15) | - |
|  | 68.9 % mild |  |
|  | 6.8 % moderate |  |
|  | 21.4 % severe |  |
| Stratum | 33 admission (32 mild, 1 moderate) |  |
|  | 28 ER (27 mild, 1 NA) |  |
|  | 42 ICU (23 mild, 6 moderate, 22 severe, 2 NA) |  |
| GOSE (interquartile range) | 7 (6-8) | - |
| Time of imaging (interquartile range) | 2 (1-16) | - |
| MRI | 62 positive/40 negative, 1 NA | - |
| CT | 54 positive/45 negative, 4 NA | - |

F, female; M, male; GCS, Glasgow Coma Score; GOSe, Glasgow Outcome Score extended; Median values are shown for the characteristics. Three stratum were present based on treatment intensity: admission to hospital, emergency room (ER visit), admission to intensive care unit (ICU).

In total there were 103 patients that had both an MRI scan and blood samples available for analysis.

A flowchart of all the analyses and data acquisition can be seen in Fig. 1.

**Figure 1.**
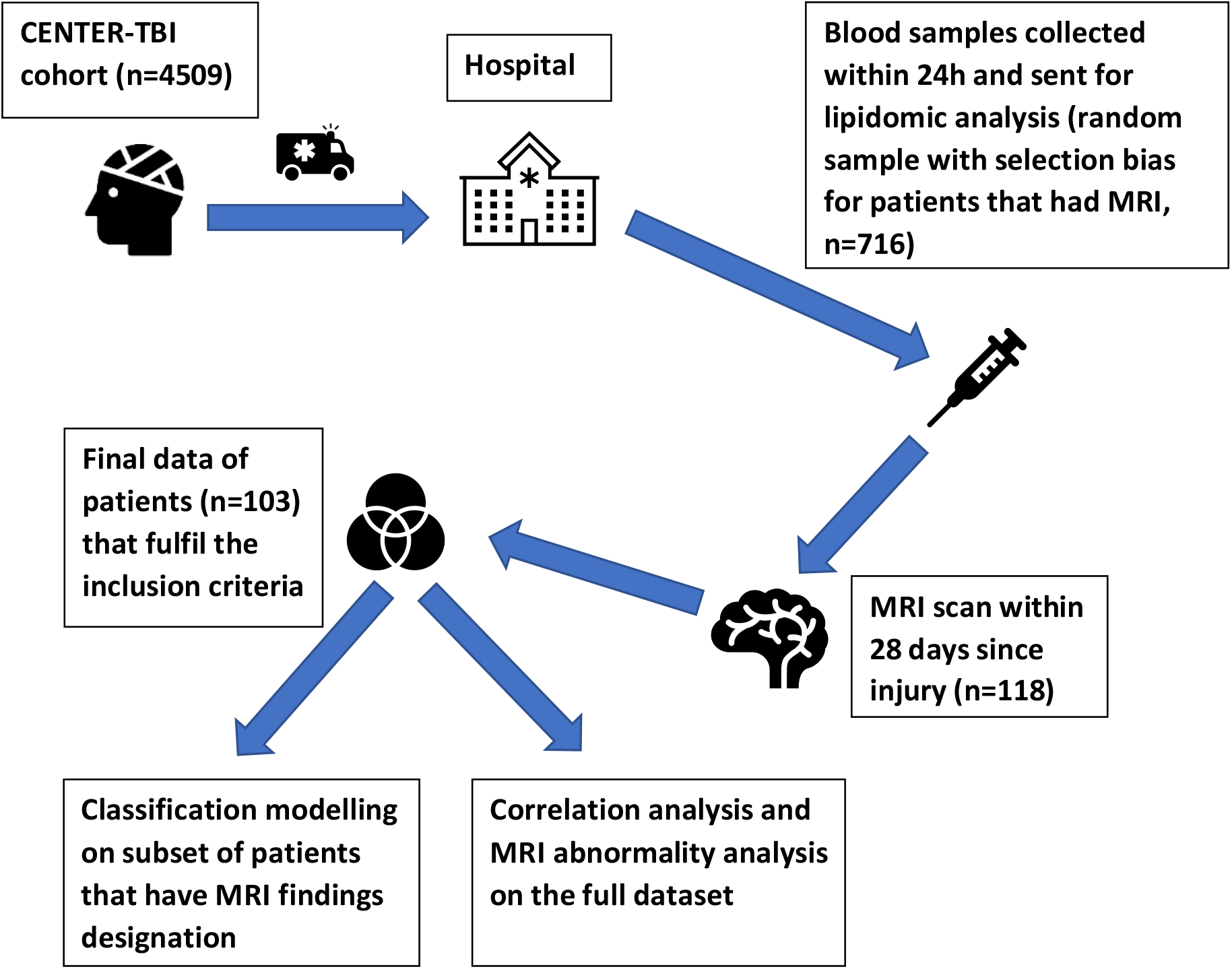
Flowchart of data acquisition and analysis. The final analysis included 103 patients that fulfilled the inclusion criteria, and the classification models included 102.

### Analysis of lipids

Blood samples of the patients were drawn within 24 hours of injury and were analyzed at Örebro University, Sweden.

Blood samples were collected using gel-separator tubes for serum and centrifuged within 60 minutes. Serum was processed, aliquoted (8 × 0·5 ml, one freeze-thaw cycle), and stored at −80 °C locally and at the central CENTER-TBI biobank (Pécs, Hungary)^18^ until shipment on dry ice to Örebro University, Sweden for analysis.

The lipidomic platform for data analysis in this study has been described in detail elsewhere^10^. The analysis was performed with an adjusted version of the Folch procedure^19^. The internal standards used, the calibration curves, the instrument description, and the sample analysis using ultra-high-performance liquid chromatography quadrupole time-of-flight mass spectrometry are the same as described previously^10^. In total, 201 known lipids were quantified that belong to the major lipidomic functional groups including ceramides (Cer), lysophosphatidylcholines (LPC), phosphatidylcholines (PC), sphingomyelins (SM), and triacylglycerols (TG).

### Image acquisition, processing, and harmonization

Images were acquired on nine 3T MRI scanners across eight sites, using study specific protocols the details of which can be found at: https://www.center-tbi.eu/project/mri-study-protocols^20^. Sequences included volumetric T1-weighted MPRAGE (voxel size 1 mm), volumetric fluid-attenuated inversion recovery, T2-weighted, susceptibility-weighted imaging and DTI. Base values of DTI were 2-mm isotropic voxels, 32 noncollinear directions, and a *b* value of 1000 seconds/mm^2^.

MRI images were reported centrally for the visible presence of lesions according to the Common Data Element (CDE) scheme for TBI (https://commondataelements.ninds.nih.gov/)^21,22^. Patients were classified as having a clinically abnormal MRI when at least one intracranial lesion secondary to TBI was detected.

All images were processed on a TBI-specific pipeline^23^. Images underwent neck cropping and were corrected for bias field inhomogeneity. Diffusion images were corrected for noise, artefacts (Gibbs, head motion and eddy currents)^24-27^ and inhomogeneities in the magnetic field^28^. Diffusion tensors were fitted via weighted least-squares to generate fractional anisotropy (FA) and mean diffusivity (MD) maps using fsl (https://fsl.fmrib.ox.ac.uk). These were non-linearly co-registered using ANTS (http://stnava.github.io/ANTs/) to the JHU-ICBM FA template^29^ to extract mean FA and MD measures for the regions of interest (Supplementary Table 2).

**Table 2.** Lipids that have the largest relative frequencies (number of correlations devided by the number of ROIs) of significant correlation to the feature sets.

| Associations with FA |  | Association with MD |  | Associations with volume |  |
| --- | --- | --- | --- | --- | --- |
| Lipid | Freq | Lipid | Freq | Lipid | Freq |
| LPC(20:4) | 0.458 | PC(O-36:4) | 0.354 | PC(36:4) | 0.431 |
| LPC(16:0) | 0.438 | PC(O-38:4) | 0.354 | TG(16:0/18:2/18:2) | 0.431 |
| LPC(20:5) | 0.396 | SM(d18:2/18:1) | 0.333 | TG(54:6) | 0.373 |
| LPC(16:0e) | 0.375 | PC(O-22:1/20:4) | 0.333 | CE fragment | 0.353 |
| LPC(18:1) | 0.375 | PC(34:3) | 0.333 | PC(O-38:6) | 0.294 |
| PC(34:3) | 0.333 | PC(O-38:5) | 0.292 | TG(18:2/22:5/16:0) | 0.216 |
| PC(35:1) | 0.292 | SM(d18:1/22:1) | 0.271 | PC(O-40:6) | 0.196 |
| PC(O-36:4) | 0.292 | SM(d41:2) | 0.271 | TG(54:6) | 0.196 |
| PC(O-38:5) | 0.292 | PC(O-40:6) | 0.229 | PC(33:0) | 0.176 |
| PC(O-40:6) | 0.292 | TG (48:4) | 0.229 | SM(d18:1/24:0) | 0.176 |
| LPC(18:2) | 0.271 | PC(P-18:0/18:1) | 0.208 | LPC(16:0) | 0.157 |
| PC(O-36:3) | 0.25 | PC(O-40:5) | 0.188 | LPC(16:0e) | 0.157 |
| PC(18:0) | 0.25 | SM(d18:1/18:1) | 0.188 | SM(d18:1/22:1) | 0.157 |
| PC(36:4) | 0.208 | PC(35:4) | 0.188 | LPC(18:0) | 0.137 |
| PC(40:8) | 0.208 | SM(d16:1/23:0) | 0.188 | LPC(20:5) | 0.137 |
| PC(O-38:4) | 0.208 | SM(d41:1) | 0.167 | PC(38:1) | 0.137 |
| SM(d36:2) | 0.188 | TG(58:10) | 0.146 | PC(O-40:5) | 0.137 |
| TG (48:4) | 0.167 | PC(36:4) | 0.146 | PC(O-40:6) | 0.137 |
| PC(35:4) | 0.146 | SM(d40:1) | 0.146 | PC(P-18:0/22:6) | 0.118 |
| PC(37:4) | 0.146 | PC(34:2) | 0.146 | SM(d18:1/24:2) | 0.118 |
| LPC(14:0) | 0.146 | PC(O-36:3) | 0.146 | LPC(22:6) | 0.098 |
| LPC(22:6) | 0.146 | SM(d18:1/24:0) | 0.146 | TG (48:4) | 0.098 |
| PC(O-34:2) | 0.146 | SM(d38:2) | 0.146 | LacCer(d18:1/16:0) | 0.078 |
| PC(O-40:5) | 0.146 | LPC(14:0) | 0.125 | LPC(18:1) | 0.078 |
| PC(P-18:0/18:2) | 0.146 | PC(35:1) | 0.125 | LPC(18:2) | 0.078 |

Differences in sites and scanners were corrected for using ComBat harmonization,^30,31^ a statistical technique to minimize unwanted scanner effect while preserving the biological variability, for the FA and MD sets (based on the healthy controls). The values of the volumetric data were normalized per patient, dividing each patients ROI values by their respective total brain volume.

For the remainder of this manuscript the 51 ROI brain volumetric segmentation will be referred to as the volumetric set, the JHU DTI-based white-matter FA map as the FA set, and the JHU DTI-based white-matter MD map as the MD set.

### Statistical analysis

All statistical analyses for this work were performed in the R statistical program version 4.1.2.

#### Frequency matrices

A correlation analysis of the frequencies of the relationships was performed for the quantitative diffusivity metrics (FA and MD) features to determine which ROI were the most frequently correlated with the lipids; and to determine if there was a topological association between the brain and the lipid concentrations. For each lipid/ROI combination the Pearson correlation was calculated together with the corresponding p-values. The correlations that were not significant after Holm correction were filtered out, through the *rcorr*.*adjust* function in the *RcmdrMisc* package. The final table had the 201 lipids by the ROIs that had significant correlations with each lipid. These remaining ROIs in the frequency table were then summed to ascertain which have the most frequent significant associations with the different lipids.

Furthermore, for each ROI, after the frequencies were calculated, it was determined if it was overall positively or negatively associated with the lipid concentrations (based on the mean value of correlations). A frequency beanplot of the ROIs was created to show the level of the associations of the ROIs with the lipids and whether their values tend to increase or decrease in relation to the concentration values of the lipids. Positive correlation is this context means that the FA and lipid values move in the same direction, *i*.*e*., both are reduced.

#### Classification models

We sought to determine whether lipidomic changes could predict whether intracranial lesions were detected on the MRI (MRI positive) or not (MRI negative) in both the full set of patients (CT positive and negative, n = 102), and as a secondary analysis in patients with no abnormalities on CT (n = 44).

A filtering process was applied to find a subset of lipids that had the highest association with the MRI findings. Two separate analyses were run, the first using a Welch t-test to find the lipids with the highest abilities to separate MRI positive/negative based on the p-values and the second based on a random forest model with the same task, and the most relevant lipids selected based on their Gini index. For each of the algorithms, the top 30 lipids associated with the MRI findings were selected, and the intersect of these two results provided the final subset of lipids for further analysis.

The subset of lipids was used as predictors in penalized logistic regression models to assess their discriminatory ability. Performance was assessed in a test set following the model fit to a training set (70%-30% split) where the subset of lipids was selected from the training set. Two penalized models were selected lasso and ridge regression, and each set of the process was repeated 100 times (data split, subset selection, model fit on training set, evaluation on the testing set), and the results of the performance for each penalized model were aggregated.

The penalized models were also run on the full set of lipids to compare the performance between them and the reduced set, and to validate the filtering process. Those analyses yielded eight under the curve values, four for the MRI positivity/negativity discrimination on the full set of patients (CT positive and negative) and four for the MRI positivity/negativity discrimination for the subset of patients with negative CT.

#### Abnormalities at the individual patient level

A separate analysis was carried on the individual patients to investigate which ones had the highest level of abnormalities defined using FA and MD. For those patients with the highest levels of abnormalities (please see below for definition) an analysis was performed to investigate whether patients who exhibited the most damage in the white matter tracts also exhibit differences in lipid concentrations either on the functional group level or the individual lipid level.

To identify which patients had the highest irregularities a comparison analysis was performed with the images of the healthy control subjects of the study. For each ROI, a linear regression model was first fit with ROI as dependent variable and sex and age (fitted as a second-degree polynomial) as predictors. The average values and a confidence internal (1 standard deviation from the baseline) for each range was calculated for the healthy controls ROI. Then, controlled for age and sex, it was determined if a patient’s ROI value was abnormal if it fell outside the baseline range. For a particular ROI, if both of the FA or MD values was outside the respective baseline range, the ROI value was designated as fully abnormal. The overall burden of injury was determined by calculating percentages of fully abnormal ROIs for each patient.

The percentage burden of injury of the patients was correlated to the 48 ROIs to see if any patterns are shown for different brain areas. For this analysis the raw concentration of the lipids within each group was added and used for a heatmap of concentration related to the burden of injury. In that plot only fully abnormal percentages are shown.

#### Abnormalities at the aggregate level

To determine if the ROIs that were prominent in the frequency of associations to the lipids were related to the the initial TBI location, that is whether the most frequent ROIs were the ones that were most affected in the injury, an abnormality frequency analysis of the ROIs was performed. In this analysis the ROIs of the TBI patients were compared with the respective ROIs of the healthy controls that undertook the scans.

For this analysis, a two-sample Kolmogorov-Smirnov test was applied for each ROI separately for the FA and MD sets to test whether the different ROIs were different between the TBI patients and healthy controls. The p-values were corrected for multiple testing with the false discovery rate method for each set separately. The frequency of significant correlations was then extracted for each ROI and these were projected on a brain map visualization using the CARIMAS software (https://turkupetcentre.fi/carimas/). For comparison, the frequency of the significant correlations of the ROIs to the lipids were also projected, as well the average value of correlation of the ROIs to the lipids.

#### Network analysis

For the construction of the partial correlation networks, a similar process as the construction of the frequency matrices was applied. In this case, the different ROIs were compared with all lipids and the lipids with the highest frequencies were found. The tables constructed in this case were 48 × 20 for the FA and MD feature sets and 51 × 20 for the volumetric set. The significant correlations between the lipids and the ROIs of the three sets were filtered out and the frequency that each lipid appears in each table was summed up. That way the lipids that correlate the most with the imaging feature sets were identified. The 40 lipids with the most correlations were then selected (20% of the total amount of lipids). The aim of this analysis was to find the lipids to be included in the partial correlation network, while the analysis of the frequency matrices was intended to visualize the patterns of relation between the ROIs and the lipids. It can point out to the ROIs that relate the most to the lipids, while the partial correlation network can show how the lipids and the brain regions are related to each other. These two analyses complement each other and elucidate the different ways that the brain regions and the circulating lipidome connect following acute TBI.

For the partial correlation network, the library *qgraph* in R was used. The top forty lipids for each feature set were used, together with the ROIs. For the FA and MD feature sets the brain was split in 3 parts, middle, left and right and the correlations were controlled with the inclusion of time elapsed between injury and blood sample draw, time between the injury and the MR scan, propofol administration and age. For the volumetric data the brain was inputted as a whole, and the relationships were controlled for age. Only the significant partial correlations are shown on the network with an alpha level of 0.05 for all sets.

## Data availability

Data are accessible based on submission of a data access request through the CENTER-TBI website: https://www.center-tbi.eu/data. CENTER-TBI is committed to data sharing, and in particular to responsible further use of the data. Hereto, we have a data sharing statement in place: https://www.center-tbi.eu/data/sharing. The CENTER-TBI Management Committee, in collaboration with the General Assembly, established the Data Sharing policy and Publication and Authorship Guidelines to assure correct and appropriate use of the data as the dataset is hugely complex and requires help of experts from the Data Curation Team or Bio-Statistical Team for correct use. This means that we encourage researchers to contact the CENTER-TBI team for any research plans and the Data Curation Team for any help in appropriate use of the data, including sharing of scripts. The complete Manual for data access is also available online: https://www.center-tbi.eu/files/SOP-Manual-DAPR-20181101.pdf

## Results

In total there were 103 patients that had both an MRI scan and blood samples available for analysis. However, one patient did not have a reliable MRI designation and was excluded from the classification model. The demographics of the study population can be seen in Table 1 and study workflow is shown in Fig. 1. In both study groups, the majority were male (TBI: 75%; control: 59%). The median age of the patients with TBI was 43 (range 18–82) years, and in healthy controls 40 years (range 21–65). The median GCS score was 15 (range 3–15) and Glasgow Outcome Scale Extended (GOSE) score 7 (range 1–8). For the baseline MRI findings there were 102 patients in the dataset that had a designation available, 62 of which were positive. For the baseline CT scan 33 were negative and a separate model only for these patients was developed (10 of 33 were MRI positive).

### Circulating lipidome associates with neuroimage findings

Overall, the results show that specific lipid classes are correlated with the findings of MRI scans. Particularly, FA tracts mostly correlated with PC and LPC, MD mostly with SM and PC, and volumetric data show most correlations to PC and TG, but other groups as well. These correlations can be seen in Table 2. In general, FA values have positive correlations to the lipids, whilst MD have negative (Supplementary Tables 3 and 4).

Overall, the patients with the highest proportions of abnormalities detected in the scans showed lower concentration of lipids, particularly in SM, Cer, and TG, consistent with previous findings^10^. Furthermore, the lipids showed a good discriminatory ability to differentiate patients who had a positive CT from those with a negative CT.

### Classification modelling for MRI/CT discrimination

The penalized models (eight in total) confirmed the discriminatory ability of the metabolites for both the MRI-positivity/negativity discrimination for all patients, and for the subset of patients with negative CT. The CT discriminatory ability was not examined in this context but only the subset of patients with negative CT. For all patients, the models had an AUC of 0.79– 0.82, whereas for the CT negative patients, the models with the reduced sets of lipids had an AUC of 0.72, and the models with the full set of lipids had an AUC of 0.6, which is likely due to the low number of observations for this analysis (Supplementary Table 5).

### Associations of lipids with ROIs

The overall lipidomic profile and the frequency of their occurrence (48 ROIs for FA and MD, 51 ROIs for volumetric data) in the correlation matrices can be seen in Table 2. The top 25 lipids for each set are shown. PC and LPC are the top lipid classes for the FA set, while SM and PC are the top lipid classes for the MD set. For the volumetric dataset, the classes that were overwhelmingly represented are PC, LPC, and TG.

The ROIs with the most relationships (frequency of at least 10%) with lipids (201 total lipids) are shown in Supplementary Tables 3, 4, and 6 as frequencies of significant correlations to the lipids. The majority of negative correlations are shown in blue and positive in red. The FA ROIs with the highest frequency of correlations to lipids are the corona radiata and the cerebellar peduncle. FA values are reduced in the TBI patients with TBI compared to the controls.

For the MD set, the corticospinal and the superior longitudinal fasciculus are the areas with the highest frequencies (Supplementary Table 3). All ROIs have negative correlations with the lipids, which means that the lipid values and the MD set values move in the opposite direction i.e. the lipid reduce and the MD values increase. For the volumetric data the ROIs overall show mixed correlations to the lipids.

Furthermore, for each ROI the values of the significant correlations were plotted as a beanplot. The correlations for the FA and the MD sets can be seen in Fig. 2. For the FA set, the correlations between ROIs and lipids are mostly positive, and for the MD set it is predominantly negative correlations, as it is for the volumetric dataset in Supplementary Fig. 3, as also shown in Supplementary Tables 3, 4, and 6.

**Figure 2.**
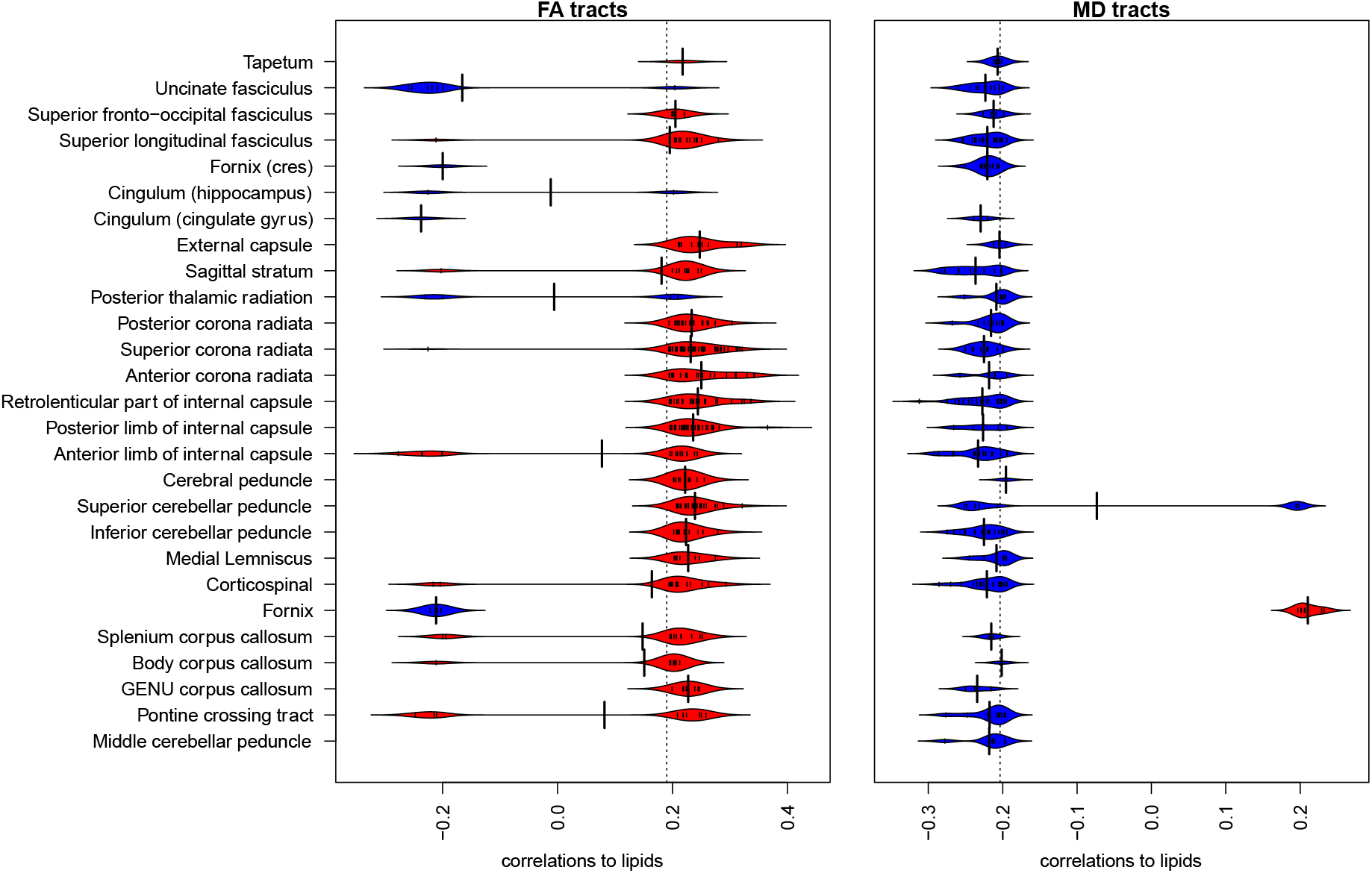
Correlations of the white matter tracts to individual lipid concentrations. The beanpots show that the fractional anisotrophy (FA) set (left) has mostly positive correlations, on average at about 0.2, while the mean diffusivty (MD) set (right) has mostly negative correlations.

**Figure 3.**
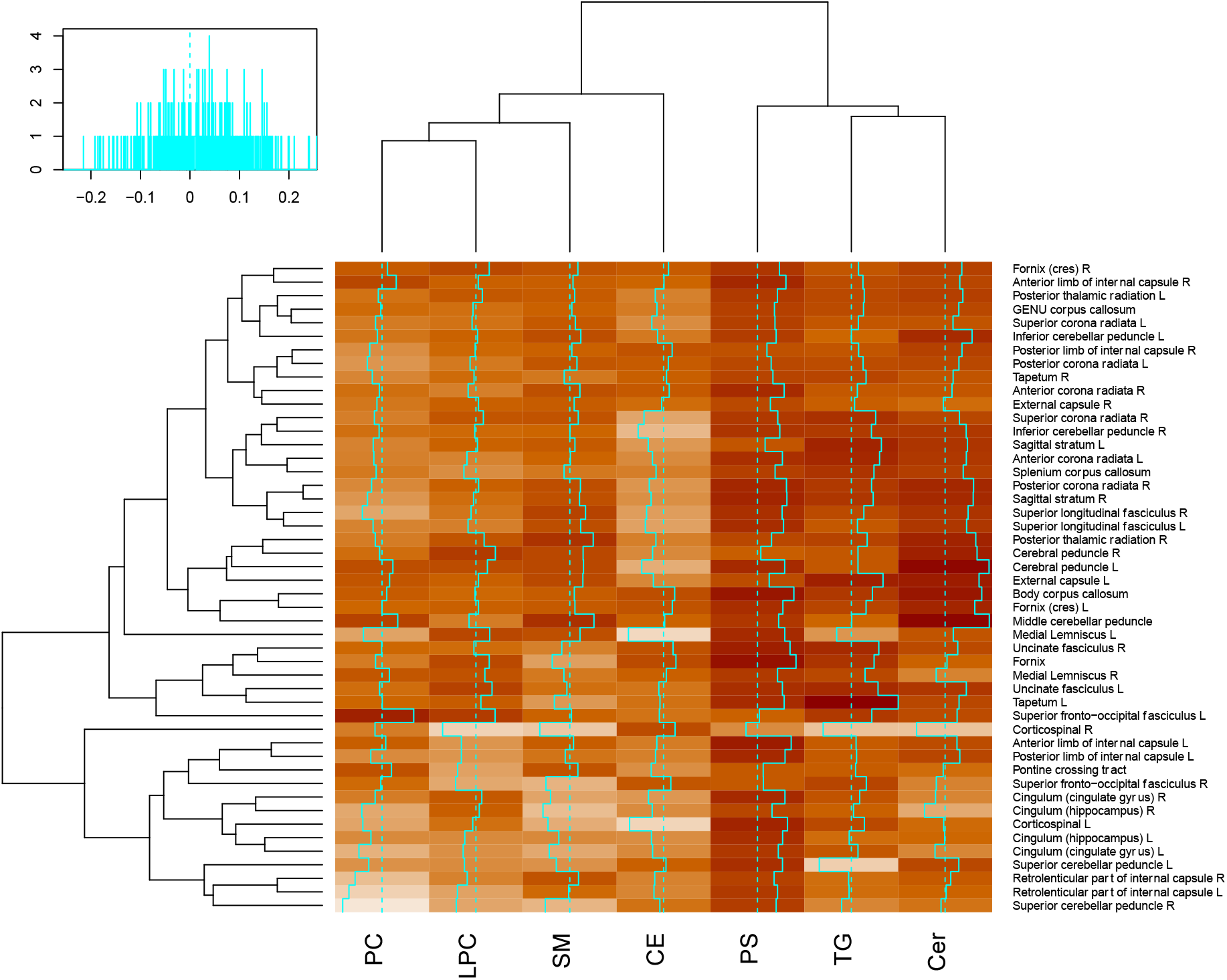
Relation of the main lipid groups to the burden of injury. The burden of injury was a summary matrix of FA and MD abonormalities, to asses the deviance from control values. The lipid group values are unadjusted summed concentrations of the lipids within each group. L, left; R, right.

The heatmap that shows the lipid values related to the burden of injury (percentages of partially and fully abnormal ROIs) is shown in Fig. 3.

### Brain maps and correlation networks of white matter tracts

In Fig. 4A, FA ROIs are mapped into the JHU brain atlas. This visualization shows the number of the abnormalities that each ROI showed in the aggregate data, the number of lipids that correlate significantly with that ROI, and the average value of these correlations. In the second row, the correlation of the lipids to the burden of injury is plotted to check if similar patterns can be seen and if the high correlations of lipids to ROIs is related to the burden of injury. Fig. 4B shows the same but for the MD set. Overall, no patterns can be seen in either figure with respect to the lipids and the frequency of abnormalities. For FA, the lipids have a positive association with the ROIs and for MD, the lipids have a negative association with the ROIs.

**Figure 4.**
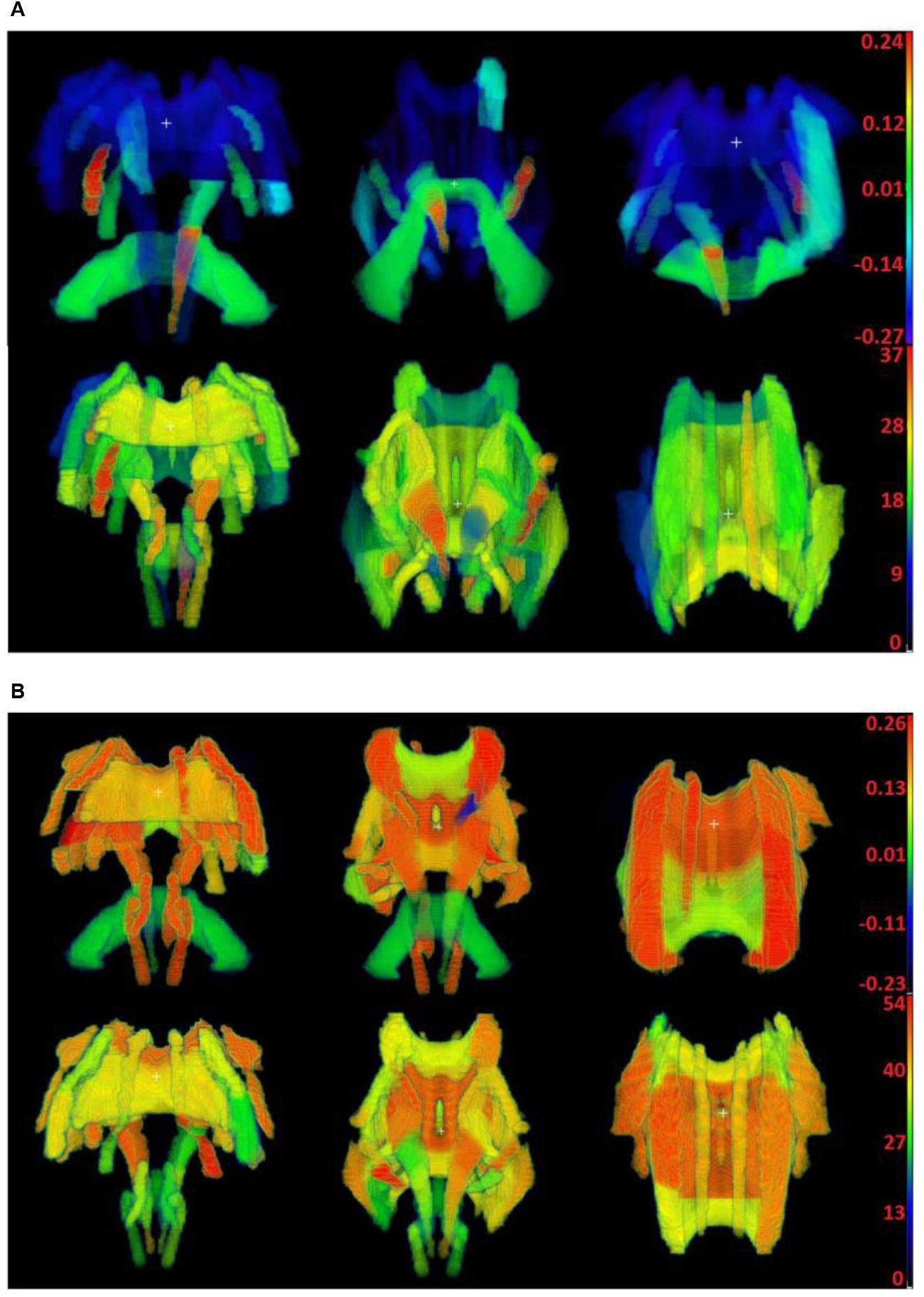
Brain maps of lipid correlations and burden of injury. On the top row of each subplot, the values of significant correlations to each brain region are shown. On the bottom row, the burden of injury for each region is shown as frequency of abnormalities per region. No visible pattern between the top and bottom rows can be seen. (**A**) FA of the white matter tracts. The correlations on top range from −0.27 (deep blue) to 0.24 (bright red) and the frequencies on bottom from 0 (black) to 37 (bright red). (**B**) MD of the white matter tracts. The correlations on top range from −0.23 (deep blue) to 0.26 (bright red) and the frequencies on bottom from 0 (black) to 54 (bright red).

The top 40 lipids that correlated with the FA set were grouped into their lipid classes, and the results are shown in Fig. 5A. PC, LPC, SM, and TG classes all show strong correlations to the different white matter tracts. For the MD set the most strongly related classes of lipids are PC and SM (Fig. 5B). Lipid levels do not seem to be affected by the time elapsed between injury and blood sample collection nor the time of scans, when controlled for in the networks.

**Figure 5.**
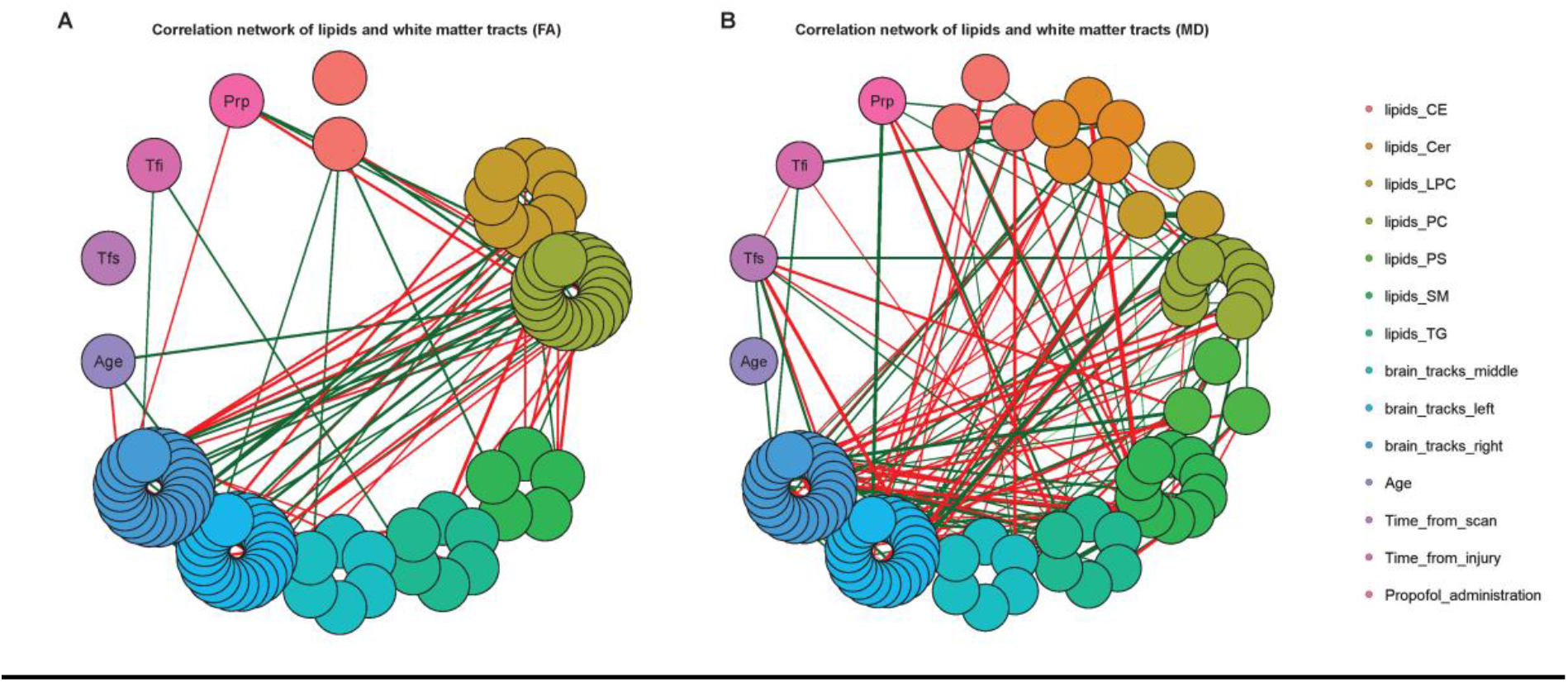
Partial correlation networks. These networks display the individual lipid correlations to other lipids, the white matter tracts, time from injury, time from scan, age, and propofol administration. The 40 lipids with the highest number of significant correlations are shown. (**A**) FA of the white matter tracts. (**B**) MD of the white matter tracts. Abbriviations: CE, cholesterol ester; Cer, ceramide;LPC, lysophosphatidylcholine; PC, phosphatidylcholine; PS, phosphatidylserine; SM, sphingomyelin; TG, triacylglycerol; FA, fractional anisotropy measures; MD, mean diffusivity.

## Discussion

In this study, we have shown that there are distinct directions of associations between the neuroimaging data (FA and MD sets) and the concentrations of circulating lipids. As expected, the FA and MD values correlated with the severity of TBI (higher MD values, lower FA)^32^. We also observed that the lipid associations to FA and MD show different metabolic signatures. LPCs associate mostly with FA while SMs associate with MD. Only PCs have strong associations with both as well as with the volumetric data.

FA has been shown to be sensitive to the changes in the microstructure of the brain^33^. However, connecting FA changes to specific brain microstructure changes is challenging due to the biologically unspecific nature of FA^34^. The primary associations with FA in our data were found with LPCs. We found previously that decreased serum LPC concentrations associated with more severe disease and poorer outcomes in TBI^10^. In animal models of TBI, LPC has been identified as one of the key lipid class that increased in the CNS following the injury^35^ and correlated with the presence of MCP-1 in the hippocampus^36^ – a key protein in attracting immune cells to the brain. However, none of these studies measured the LPCs concentrations in the blood. There are known transporters of LPCs in the CNS^37^ and decreased levels of circulating LPC have been associated with poor outcomes in other CNS diseases^38^. LPCs are also implicated in a wide range of inflammatory diseases^39^. Given these previous findings, the positive associations of LPCs with FA as observed in our study may reflect an increased inflammatory drive associating with changes in brain microstructure following a TBI.

SM is a key lipid class needed for the synthesis of myelin in the CNS^40^. Decreased serum levels of SMs following a TBI and inverse associations with MD changes suggest that SMs are recruited for myelin repair following the membrane damage in TBI. Similar inverse relationship between SMs levels and MD has been observed in Alzheimer’s disease^41^. SMs are also converted to ceramide in inflammatory conditions, which could be another non-exclusive cause for reduced levels of circulating SMs.

The lipids, which were consistently associated both with the diffusion imaging and the volumetric data were PCs. PCs are a key component of cellular membranes and therefore changes in brain microstructure will disrupt PC metabolism^42^. PCs can also be broken down into free fatty acids, which can be utilized as an energy source in the CNS. Other acute brain injuries have been shown to increase activity of enzymes involved in fatty acid metabolism immediately following the injury^43^. We have earlier shown that medium-chain fatty acids are associated with severity of TBI^9,10^. It is known that TBI causes an energy crisis within the CNS^44^ and therefore the increased utilization of PCs could be related to the severity of the energy crisis that is reflected in the poor imaging metrics.

The association with quantitative white matter imaging metrics and PCs was global as can be seen in Fig. 3. This global distribution of association is seen to a lesser extent with the TGs. In contrast, the ceramide associations seem to form two main clusters; one of more central fibres with the strongest associations and the other of the longer tracts with lower association. CEs and PCs associations were also stronger with these longer white tracts, while SMs were associated with more central tracts. The pathophysiology of what is leading to theses different predilections for different types and regions of white matter is likely to be complex. An increase in ceramide in the plasma has been associated with several inflammatory and other neurodegenerative diseases including multiple scleroisis^45^and Alzheimer’s disease^46^. The different roles CEs and ceramides play in emergy balance may influence different patterns seen^47^.

The observed lack of lipid associations with specific brain volume regions in the gray matter suggests that changes in the levels of circulating lipids do not reflect the primary cortical injury in TBI. However, the consistant associations with volumetric data from with deeper white matter structures suggest that the lipid changes reflect this deeper injury type. This could result from the secondary brain injury driven by axonal damage occurring in these structures. However, these findings are in line with our previous work, where we could see clear assocations between polar metabolites and deeper brain volume changes^48^. This suggests that the patterns of associations between the volumes and metabolites regardless of type is more clear in the deeper brain regions.

## Limitations

This is the largest study to date combining serum metabolomics and MRI imaging including the diffusion-weighted imaging. However, the sample size is still small and further validation studies are required to support our findings. The underlying reasons for the lipid changes observed are also difficult to pinpoint due to the difficulty in obtaining tissue or fluid samples from within the CNS, and therefore studies in suitable experimental models would be needed in order to understand the causes of the observed lipid changes.

## Conclusions

We have identified groups of lipids which assocate with specific MRI imaging metrics following TBI. There appears to be consistent patterns of lipid changes associating with the specific microstructure changes in the CNS white matter. There is also a pattern of lipids with regional specficity, suggesting that blood-based lipidomics may provide an insight into the underlying disease mechanisms in TBI.

## Abbreviations

CE: cholesterol ester
Cer: ceramide
CT: computed tomography
FA: fractional anisotropy measures
LPC: lysophosphatidylcholine
MD: mean diffusivity
MS: mass spectrometry
PC: phosphatidylcholine
PS: phosphatidylserine
SM: sphingomyelin
TBI: traumatic brain injury
TG: triaclyglycerol

## Acknowledgements

The authors thank Daniel Duberg, Lisanna Sinisalu, and Vanja Stefanovic for technical assistance in metabolomics analysis.

## Supplementary material

Supplementary Tables 1-6

Supplementary Figure 1

