## Supplementary Material for "Serum lipidome associates with neuroimaging features in patients with traumatic brain injury"

Ilias Thomas et al.

### Supplementary Material

**Supplementary Table 1.** The volumetric data ROIs as segmented into 51 regions.

| ROI Index | Collapsed ROI name |
|---|---|
| 1 | BrainStem |
| 2 | CerebellarVermis |
| 3 | LeftAcumbens |
| 4 | LeftAmygdala |
| 5 | LeftBasalForebrain |
| 6 | LeftCaudate |
| 7 | LeftCerebellarWhite |
| 8 | LeftCerebellumGrey |
| 9 | LeftCerebralWhiteMatter |
| 10 | LeftDorsolateralFrontal |
| 11 | LeftFrontalPole |
| 12 | LeftFusiformGyrus |
| 13 | LeftHippocampalComplex |
| 14 | LeftInsula |
| 15 | LeftLateralParietalLobe |
| 16 | LeftMedialFrontal |
| 17 | LeftMedialParietalLobe |
| 18 | LeftOccipitalLobe |
| 19 | LeftOrbitofrontal |
| 20 | LeftPallidum |
| 21 | LeftPrecentral |
| 22 | LeftPutamen |
| 23 | LeftSubcallosal |
| 24 | LeftTemporalLobe |
| 25 | LeftTemporalPole |
| 26 | LeftThalamus |
| 27 | RightAcumbens |
| 28 | RightAmygdala |
| 29 | RightBasalForebrain |
| 30 | RightCaudate |
| 31 | RightCerebellarWhite |
| 32 | RightCerebellumGrey |

| | |
|---|---|
| 33 | RightCerebralWhiteMatter |
| 34 | RightDorsolateralFrontal |
| 35 | RightFrontalPole |
| 36 | RightFusiformGyrus |
| 37 | RightHippocampalComplex |
| 38 | RightInsula |
| 39 | RightLateralParietalLobe |
| 40 | RightMedialFrontal |
| 41 | RightMedialParietalLobe |
| 42 | RightOccipitalLobe |
| 43 | RightOrbitofrontal |
| 44 | RightPallidum |
| 45 | RightPrecentral |
| 46 | RightPutamen |
| 47 | RightSubcallosal |
| 48 | RightTemporalLobe |
| 49 | RightTemporalPole |
| 50 | RightThalamus |
| 51 | Ventricle |

**Supplementary Table 2.** The white matter track data as segmented into 48 regions ROIs.

| **ROI index** | **FA and MD ROIs** |
|---|---|
| 1 | Middle cerebellar peduncle |
| 2 | Pontine crossing tract (a part of MCP) |
| 3 | Genu of corpus callosum |
| 4 | Body of corpus callosum |
| 5 | Splenium of corpus callosum |
| 6 | Fornix (column and body of fornix) |
| 7 | Corticospinal tract R |
| 8 | Corticospinal tract L |
| 9 | Medial lemniscus R |
| 10 | Medial lemniscus L |
| 11 | Inferior cerebellar peduncle R |
| 12 | Inferior cerebellar peduncle L |
| 13 | Superior cerebellar peduncle R |
| 14 | Superior cerebellar peduncle L |
| 15 | Cerebral peduncle R |
| 16 | Cerebral peduncle L |
| 17 | Anterior limb of internal capsule R |
| 18 | Inferior cerebellar peduncle L |
| 19 | Posterior limb of internal capsule R |
| 20 | Posterior limb of internal capsule L |
| 21 | Retrolenticular part of internal capsule R |
| 22 | Retrolenticular part of internal capsule L |
| 23 | Anterior corona radiata R |
| 24 | Anterior corona radiata L |
| 25 | Superior corona radiata R |
| 26 | Superior corona radiata L |
| 27 | Posterior corona radiata R |
| 28 | Posterior corona radiata L |
| 29 | Posterior thalamic radiation (include optic radiation) R |
| 30 | Posterior thalamic radiation (include optic radiation) L |
| 31 | Sagittal stratum (include inferior longitidinal fasciculus and inferior fronto-occipital fasciculus) R |

| | |
|---|---|
| 32 | Sagittal stratum (include inferior longitidinal fasciculus and inferior fronto-occipital fasciculus) L |
| 33 | External capsule R |
| 34 | External capsule L |
| 35 | Cingulum (cingulate gyrus) R |
| 36 | Cingulum (cingulate gyrus) L |
| 37 | Cingulum (hippocampus) R |
| 38 | Cingulum (hippocampus) L |
| 39 | Fornix (cres) / Stria terminalis (can not be resolved with current resolution) R |
| 40 | Fornix (cres) / Stria terminalis (can not be resolved with current resolution) L |
| 41 | Superior longitudinal fasciculus R |
| 42 | Superior longitudinal fasciculus L |
| 43 | Superior fronto-occipital fasciculus (could be a part of anterior internal capsule) R |
| 44 | Superior fronto-occipital fasciculus (could be a part of anterior internal capsule) L |
| 45 | Uncinate fasciculus R |
| 46 | Uncinate fasciculus L |
| 47 | Tapetum R |
| 48 | Tapetum L |

**Supplementary Table 3.** The FA ROIs that show the most correlations to the lipids together with the frequencies and the sign of the average correlations.

| Frequency | Sign of correlation | Name |
|---|---|---|
| 0.323 | + (positive) | Superior corona radiata L |
| 0.209 | + (positive) | Superior cerebellar peduncle R |
| 0.204 | + (positive) | Medial Lemniscus L |
| 0.194 | + (positive) | Corticospinal L |
| 0.184 | + (positive) | Superior longitudinal fasciculus L |
| 0.159 | + (positive) | Posterior limb of internal capsule L |
| 0.159 | + (positive) | Superior corona radiata R |
| 0.154 | + (positive) | Retrolenticular part of internal capsule R |
| 0.149 | + (positive) | Cerebral peduncle L |
| 0.139 | + (positive) | Posterior limb of internal capsule R |
| 0.124 | + (positive) | Inferior cerebellar peduncle L |
| 0.124 | + (positive) | Posterior corona radiata L |
| 0.114 | + (positive) | Anterior corona radiata L |

**Supplementary Table 4.** The MD ROIs that show the most correlations to the lipids together with the frequencies and the sign of the average correlations.

| Frequency | Sign of correlation | Name |
|---|---|---|
| 0.184 | - (negative) | Corticospinal L |
| 0.154 | - (negative) | Fornix (cres) L |
| 0.119 | - (negative) | Superior longitudinal fasciculus L |
| 0.1 | - (negative) | Retrolenticular part of internal capsule R |

**Supplementary Table 5.** Findings of the MRI classification performance of the lipids. The results of the ridge reduced model are shown.

| | (1/0/NA) | AUC (CI) |
|---|---|---|
| MRI findings (n=102) | (62/40) | 0.85 (0.71- 0.98) |
| Subset of patients with negative CT (n=45) | (11/33/1) | 0.7 (0.37- 0.98) |

**Supplementary Table 6.** The volumetric ROIs that show the most correlations to the lipids together with the frequencies and the sign of the average correlations.

| Frequency | Sign of correlation | Name |
|---|---|---|
| 0.159 | + (positive) | RightAcumbens |
| 0.129 | - (negative) | LeftMedialParietalLobe |
| 0.129 | + (positive) | RightSubcallosal |
| 0.119 | + (positive) | LeftBasalForebrain |
| 0.119 | - (negative) | LeftFusiformGyrus |
| 0.114 | - (negative) | RightBasalForebrain |
| 0.109 | + (positive) | LeftSubcallosal |
| 0.104 | + (positive) | LeftFrontalPole |

**Supplementary Figure 1.** Beanpot of the correlation values of the volumetric ROIs to all the lipids. Predominantly negative correlations are seen which, on average, are at about -0.2.

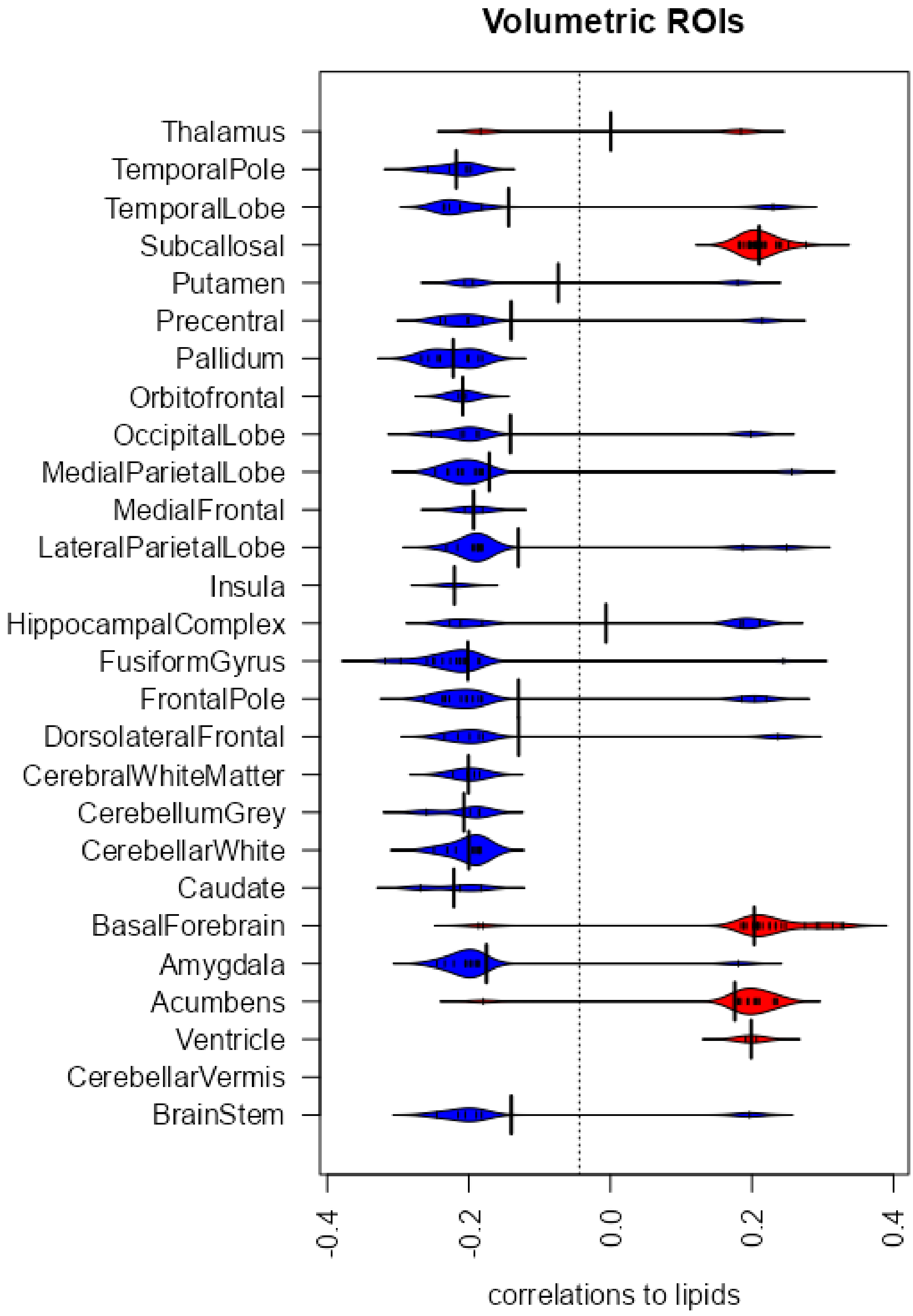
